# Joint-Label Fusion Brain Atlases for Dementia Research in Down Syndrome

**DOI:** 10.1101/2020.11.10.20228742

**Authors:** Nazek Queder, Michael J. Phelan, Lisa Taylor, Nicholas Tustison, Eric Doran, Christy Hom, Dana Nguyen, Florence Lai, Margaret Pulsifer, Julie Price, William C. Kreisl, Diana H. Rosas, Sharon Krinsky-McHale, Adam Brickman, Michael A. Yassa, Nicole Schupf, Wayne Silverman, Ira T. Lott, David B. Keator

**Author notes:** Corresponding Author: Nazek Queder, University of California, Irvine, Irvine Hall rm. 164 - Zot 3960, Irvine, CA 92697.

## Abstract

Research suggests a link between Alzheimer’s Disease in Down Syndrome (DS) and the overexpression of amyloid plaques. Using Positron Emission Tomography (PET) we can assess the in-vivo regional amyloid load using several available ligands. To measure amyloid distributions in specific brain regions, a brain atlas is used. A popular method of creating a brain atlas is to segment a participant’s structural Magnetic Resonance Imaging (MRI) scan. Acquiring an MRI is often challenging in intellectually-imparied populations because of contraindications or data exclusion due to significant motion artifacts or incomplete sequences related to general discomfort. When an MRI cannot be acquired, it is typically replaced with a standardized brain atlas derived from neurotypical populations which may be inappropriate for use in DS. In this project, we create a series of disease and diagnosis-specific (cognitively stable, mild cognitive impairment (MCI-DS), and dementia) probabilistic group atlases of participants with DS and evaluate their accuracy of quantifying regional amyloid load compared to our ground truth individual MRI-based segmentations. Further, we compare the diagnostic-specific atlases with a probabilistic atlas constructed from similar-aged cognitively-stable neurotypical participants. We hypothesized that regional PET signals will best match the ground truth by using DS group atlases that aligns with a participant’s disorder and disease status (e.g. DS and MCI-DS). Our results vary by brain region but generally show that using a disorder-specific atlas in DS better matches the ground truth than using an atlas constructed from cognitively-stable neurotypical participants. We found no additional benefit of using a disease-state specific atlas. All atlases are made publicly available for the research community.

**Abbreviations:** AD, DS, Aβ, DSG, CS-DS, CS-NT, LOOCV, ROI, MSE, MRI, PET, JLF, CS, MCI-DS, DEM.

## 1. INTRODUCTION

Down Syndrome (DS), a genetic disorder caused by abnormal cell division resulting in a full or partial extra copy of chromosome 21, is the most common chromosomal disorder with an incidence rate of approximately 1 in every 700 babies born (Parker et al., 2010). Trisomy 21 impacts neurological development in DS leading to phenotypic abnormalities and cognitive deterioration, affecting memory and intellectual abilities (Gardiner et al., 2010; Silverman, 2007). Further, developmental abnormalities impact brain region growth, resulting in brain morphometry that is, on average, different from neurotypicals’ (Lee et al., 2016).

Unlike other populations, individuals with DS have distinctive brain characteristics and craniofacial phenotypes including microcephaly, platybasia, increased biparietal to occipitofrontal diameter ratio, and approximately 20% reduction in brain volume compared to age-matched controls (Rodrigues et al., 2019). Stereological measurements of Magnetic Resonance Imaging (MRI) suggest an overall reduction in the cerebral and cerebellar hemispheres in individuals with DS compared to healthy age-matched neurotypicals (Raz et al., 1995; Rodrigues et al., 2019; Weis et al., 1991). Individuals with DS and dementia have MRI findings that are comparable to AD patients’ (Prasher et al., 2003; Radhakrishnan and Towbin, 2014) including diffuse cortical atrophy in the parietal lobes, broadening of the marginal branch of the cingulate sulcus, central, post-central, intraparietal, parieto-occipital sulcus, entorhinal cortex, amygdala, and hippocampus (Ballard et al., 2016; Lai and Williams, 1989). Additional MRI evidence shows that after correcting for total brain volume, there is a significant reduction in total brain, gray matter, ventricles, and left amygdala and hippocampus in participants with DS compared to healthy neurotypicals and there’s even more reduction in those with dementia and DS when compared to healthy controls (Pearlson et al., 1998). Further, Computerized Tomography (CT) findings show increased atrophy in participants with DS and dementia compared to those without dementia (Devinsky et al., 1990; LeMay and Alvarez, 1990; Lott and Lai, 1982).

Positron Emission Tomography (PET) has been used to identify amyloid deposition in the brain and is a widely used neuroimaging tool for Alzheimer’s Disease (AD) research (Quigley et al., 2011; Rabinovici and Jagust, 2009). Individuals with DS are at a higher risk of developing neuropathological phenotypes that are indistinguishable from AD; about fifty percent of the population develop dementia at age 40 and seventy percent at age 60 and up (Lott & Head, 2001; SCHAPIRO and B, 1986; Schapiro et al., 1992). The early progression of AD in participants with DS is thought to be associated with the expression of amyloid-beta (Aβ) (Doran et al., 2017; Moncaster et al., 2010) and thus measuring brain Aβ accurately may help to identify those at risk of early transition to dementia (Keator et al., 2020a). Due to the lack of structural information in amyloid PET scans, researchers investigating regional Aβ burden typically obtain an MRI scan to evaluate amyloid distribution and identify regional anatomic boundaries, which serves as a brain atlas. However, researchers face many challenges in acquiring MRI scans of intellectually-imparied populations (Ali et al., 2013; Brockmeyer, 1999; Wullink et al., 2009). Often the acquisition of an MRI scan is not possible in these populations because of contraindications or the data is unusable due to significant motion artifacts, incomplete sequences related to claustrophobia and/or general discomfort, or related problems. When an MRI is not available, researchers use one of several canonical brain atlases, often constructed from a single, young, healthy individual, or groups of individuals, whose brain morphometry is vastly different from those with DS. Such atlases may not accurately reflect DS morphology thus result in inaccurate quantification of regional amyloid binding.

Neuroimaging research in DS provides evidence of distinctive structural characteristics in addition to other risk factors, like amyloid expression, that are thought to be linked to the early progression of AD like dementia (Annus et al., 2016; Cole et al., 2017; Mann et al., 1990). Investigations of AD in DS rely on our ability to accurately detect Aβ in implicated regions of the brain which provides understanding of amyloid distribution in the development of dementia both efficiently and effectively. Given the challenges researchers face in structural brain mapping and the complexity and variation in atrophy, shape, and size of the human brain, accurate structural brain templates that account for such variations and represent the population accurately are required (Toga and Thompson, 2002).

In this manuscript, we created a series of probabilistic disease-state (i.e. cognitively stable (CS), mild cognitive impairment (MCI-DS), or dementia (DEM)) and disorder-specific (i.e. DS) brain atlases from participants with DS. We evaluated the accuracy of using these group-based atlases to compute regional amyloid distributions by disease status compared to using individualized MRI-based segmentations (IM) or using a group atlas constructed from similar-aged cognitively-stable neurotypical participants. We hypothesized that using group atlases which match the disease state and disorder will result in regional amyloid measurements that are most sensitive to disease status and most consistent with the ground-truth individualized MRI-based segmentations.

## 2. METHODS

### 2.1 Participants

Neuroimaging data of participants with DS were collected as part of the Alzheimer’s Biomarkers Consortium-Down Syndrome (ABC-DS) and neurotypicals from the Neuroimaging Biomarkers for Cognitive Decline in Elderly with Amyloid Pathology (NIA) project at UCI. Data from 83 participants with DS (Table 1) were collected at three participating sites: Massachusetts General Hospital/Harvard University (MGH), Columbia University, and University of California, Irvine (UCI). The sample included participants with three consensus diagnoses: cognitively stable (CS) - participants who show no signs of dementia or cognitive decline; mild cognitive impairment (MCI-DS) - participants with DS who show mild cognitive detorioration but do not satistfy the criteria for dementia; demented (DEM) - participants with DS showing severe cognitive impairment (Silverman et al., 2004). The neurotypical dataset from the NIA study consisted of 56 participants collected at UCI.

**Table 1:**
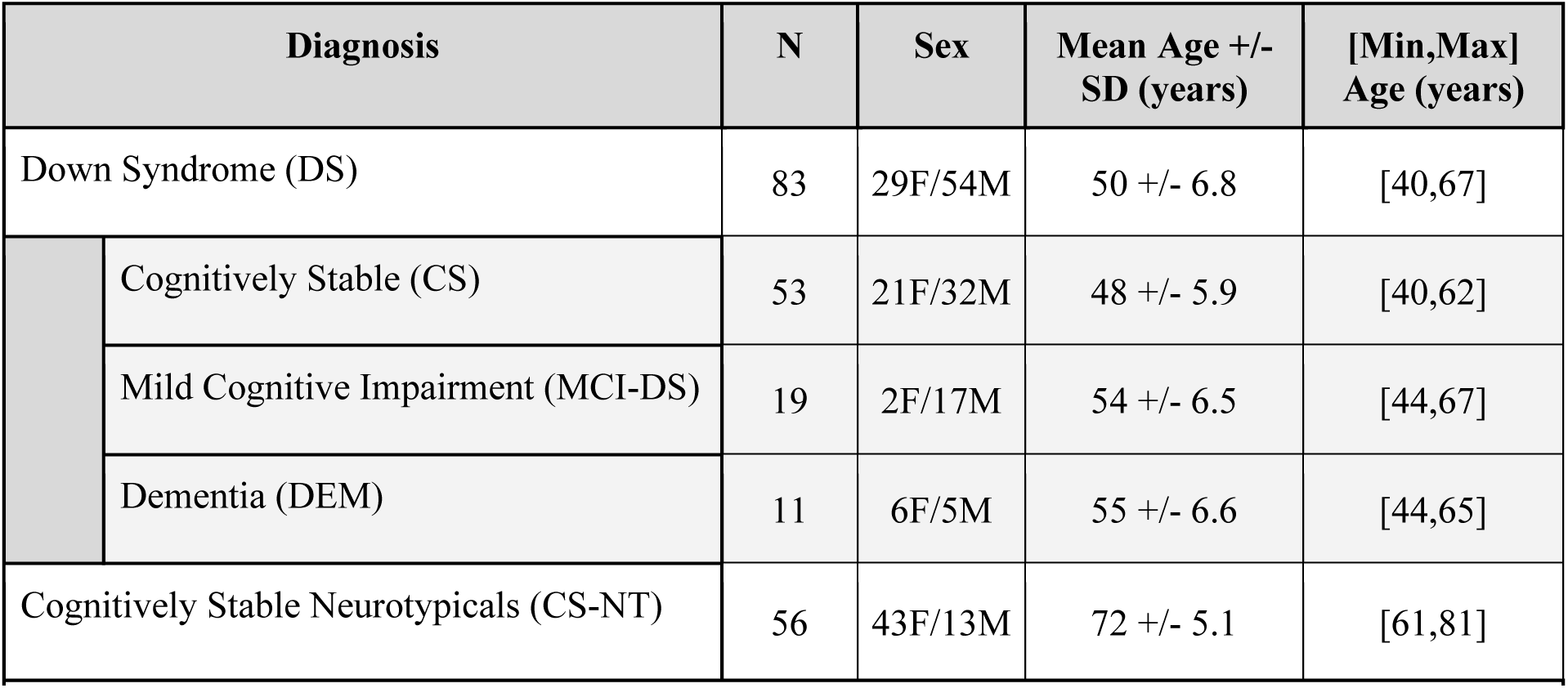
Participant characteristics showing the total sample size (N), sex, age mean +/-standard deviation, and minimum/maximum age in years.

| Diagnosis |  | N | Sex | Mean Age +/- SD (years) | [Min,Max] Age (years) |
| --- | --- | --- | --- | --- | --- |
| Down Syndrome (DS) |  | 83 | 29F/54M | 50 +/- 6.8 | [40,67] |
|  | Cognitively Stable (CS) | 53 | 21F/32M | 48 +/- 5.9 | [40,62] |
|  | Mild Cognitive Impairment (MCI-DS) | 19 | 2F/17M | 54 +/- 6.5 | [44,67] |
|  | Dementia (DEM) | 11 | 6F/5M | 55 +/- 6.6 | [44,65] |
| Cognitively Stable Neurotypicals (CS-NT) |  | 56 | 43F/13M | 72 +/- 5.1 | [61,81] |

### 2.2 Image Acquisition

T1-weighted MRI scans for the DS sample acquired at UCI were scanned on a Philips Achieva 3T (orientation=sagittal, TR/TE=7.8/3.6ms, flip angle=7°,voxel size= 1 mm^3^, matrix size=256×256×176) or a Siemens Prisma 3T (orientation=sagittal, IT/TR/TE=900/2300/3.0ms, flip angle=9°, voxel size=1 mm^3^, matrix size=240×256×208). Both Columbia and MGH used a Siemens Prisma 3T scanner with consistent protocols. For the NIA study, participants were scanned at UCI on a Siemens Prisma 3T (orientation=coronal, IT/TR/TE=902/2300/2.38ms, flip angle=8°, voxel size=0.8 mm^3^, matrix size=320×320×240).

PET scan data for the DS sample from UCI was acquired on a High Resolution Research Tomograph (HRRT; orientation=axial, voxel size=1.2mm^3^, matrix size=256×256×207, reconstruction=OP-OSEM3D), at Columbia on a Siemens Biograph64 mCT (orientation=axial, voxel size=1.0×1.0×2.0mm, matrix size=400×400×436, reconstruction=OSEM3D+TOF 4i21s), and MGH on a Siemens Biograph mMR (orientation=axial, voxel size=2.1×2.1×2.0mm, matrix size=344×344×127, reconstruction=OP-OSEM 3i21s). Image acquisition followed the Alzheimer’s Disease Neuroimaging Initiative (ADNI) (“ADNI Home,” n.d.; Landau et al., 2012) protocol: 4×5 minute frames collected 50-70 minutes after injection. PET reconstructions were performed with all corrections as implemented on each platform. PET scan processing followed the methods described in Keator et al. (Keator et al., 2020b), consisting of realignment and averaging of the PET frames, coregistration with the structural MRI, and standardized uptake value ratio (SUVR) scaling using the each participant’s cerebellum cortex reference region.

### 2.2 Atlas Creation

In this study, we created a series of high resolution brain atlases using the Advanced Normalization Tool’s (ANTs version 2.2.0; (Avants et al., 2011); RRID:SCR_004757) joint label fusion (JLF) algorithm. The JLF atlases were created from individual MRI-based segmentations computed with the FreeSurfer (FS6 version 6.0; RRID:SCR_001847)(Fischl et al., 2004, 2002; “FreeSurfer,” n.d.) tool and the Desikan/Killiany atlas (Desikan et al., 2006). After the initial Freesurfer reconstruction, all segmentations were visually checked for accuracy, and corrected when necessary using procedures from FreeSurfer. The resulting volumetric version of the aparc+aseg segmentations from Freesurfer and the associated T1-weighted structural images were used in creating the JLF atlases on the UCI High Performance Computing cluster. To align the PET scan images with the JLF atlases, ANTs symmetric diffeomorphic image registration algorithm was used to register each participant’s structural MRI to each JLF atlas and apply the resulting deformations to the coregistered PET scan. PET scan region of interest (ROI) average SUVR values were then extracted from each JLF atlas and from the individual Freesurfer segmentations (IM) for the comparisons presented in section 3. The following regions were chosen based on existing AD literature implicating them in the progression of disease: inferior parietal, lateral occipital, superior frontal, rostral medial frontal, medial/inferior temporal, anterior cingulate, superior temporal, dorsal striatum, middle temporal, posterior cingulate, medial/lateral orbitofrontal, hippocampus, and entorhinal cortex.

#### 2.2.1 Disorder and Disease-State-Specific Group (DSG) Atlas

To evaluate the accuracy of using a JLF group atlas within-disorder and consistent with a participant’s disease status (e.g. CS, MCI-DS, DEM), a series of atlases were constructed. DSG atlases were constructed using Freesurfer IM segmentations separately for the three main consensus diagnoses reflecting DS participants’ dementia status: CS, MCI-DS, and DEM. For the PET comparisons, we created many DSG atlases, leaving one participant out (LOOCV) in each fold, to compare with their IM-based PET region-of-interest (ROI) averages. This was done to prevent bias in the JLF-derived regional PET measures as would be the case if the participant’s data being evaluated was used in construction of the JLF atlas.

#### 2.2.2 Disorder-Specific Cognitively-Stable Down Syndrome (CS-DS) Group Atlas

In section 2.2.1 we constructed group atlases which took into account the disease status in Down Syndrome (DSG atlas). Unlike the DSG atlas, here we build an atlas of the aged brain of participants with DS who were cognitively stable (CS-DS), to evaluate whether accounting for disease status in a group atlas is necessary. To evaluate the CS-DS atlas, we compared the IM PET ROI averages in participants who had either a consensus-based diagnosis of MCI-DS or DEM and thus did not need to use the LOOCV paradigm as described for the DSG atlas comparisons because the atlas and comparison groups were independent.

#### 2.2.3 Cognitively-Stable Neurotypical (CS-NT) Group Atlas

In sections 2.2.1 and 2.2.2 we evaluated group atlases which take into account the disease status and disorder. Here we construct a group atlas from the cognitively-stable neurotypical (CS-NT) participants to evaluate whether accounting for the disorder (e.g. DS) or disease status is necessary for accurately reproducing the PET ROI averages calculated using the ground-truth individual IM segmentations. Similar to the CS-DS atlas, we build an atlas reflecting the aged brain (72 +/-5.1 years) of participants without DS who were cognitively stable. For evaluations of the CS-NT atlas, we compare the IM PET ROI averages in each of the diagnostic groups with DS (i.e. CS, MCI-DS, and DEM). Because the CS-NT group is independent from each of the DS groups, the LOOCV paradigm was not needed.

### 2.3 PET ROI Analysis

To quantify differences in amyloid average values between the three diagnostic groups in our ROIs of interest, we built fixed-effects linear regression models for each ROI separately using RStudio (version 1.1447; RRID:SCR_001905). The dependant variable was the difference in amyloid load between our atlases (i.e. DSG, CS-DS, CS-NT) and the individual freesurfer segmentations (i.e. IM) as a function of the independent variables: average amyloid values between the two atlases, diagnosis, and the interaction between the average amyloid load and the diagnosis. Results (section 3) are illustrated using Bland Altman plots (Bland et al., 1986); (Cardemil, 2017).

## 3. RESULTS

### 3.1 Atlas Structure Comparisons

To quantify the overall structural differences between the atlases, we computed mean squared error (MSE) between the DSG (disease-state-specific - Down Syndrome), CS-DS (cognitively stable - Down Syndrome), and CS-NT (cognitively stable - neurotypical) atlases compared to the individual Freesurfer segmentations (IM). MSE scores were normalized to the MSE score of comparing the JLF atlas to a matrix of zeros, scaled to percentage, and compared across the atlases for each diagnostic group (section 3.1.1) using one-way analysis of variance (ANOVA) and Tukey’s honestly significant difference (TukeyHSD) testing in RStudio (version 1.1447; RRID:SCR_001905). *P*-values from the overall ANOVA were adjusted using the Bonferroni-Hommel method (Hommel, 1988). Box plots of the results are shown in Figures 2 and 3. A visual example of the DSG, CS-DS, and CS-NT atlases compared to a selected IM atlas is included in the supplemental material (S1).

**Figure 2:**
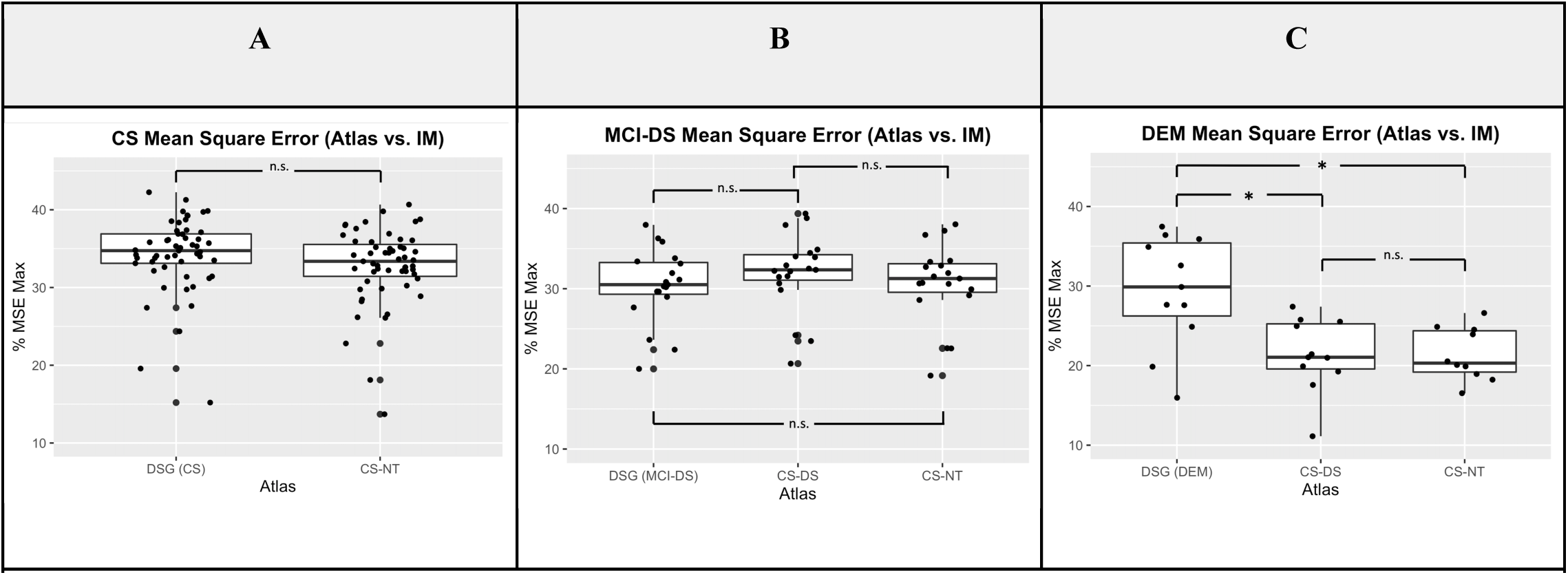
Mean Square Error (MSE) by Atlas. Higher percentage of MSE score indicates worse match between our atlas and the individual MRI-based Freesurfer segmentations (IM). Asterisks identify results that are significant at p<0.05 after adjusting for the p-value, whereas “n.s.” indicates non-significant.

#### 3.1.1 Comparisons Across Atlases within Diagnostic Groups

Here we evaluate the overall structural difference in atlas labels between the Freesurfer segmentations and each group atlas across the diagnostic groups. The overall ANOVA models of the CS (Figure 2, panel A) and MCI-DS (Figure 2, panel B) diagnostic groups were not significantly different across the atlases. In comparison, the DEM overall model was significant across the atlases (*F*(2,30) = 8.669, *p*_*adj*._*<*.003; Figure 2, panel C). A post hoc Tukey test showed that, in the DEM group, both the CS-DS and the CS-NT atlases had a mean MSE score that was significantly different from the DSG atlas (CS-DS vs. DSG (*t*(20) = −8.013, *p*_*adj*._ < .006); CS-NT vs. DSG (*t*(20) = −9.001, *p*_*adj*._ < .002)). While no significant differences were observed when comparing the CS-DS and the CS-NT atlases.

In summary, both the CS and MCI-DS groups’ average MSE scores remained consistent across each of the atlases, indicating that structural differences between the group atlases and the Freesurfer individual segmentations (IM) are likely insignificant. Interestingly, in the DEM group, the DSG atlas had a significantly higher mean MSE score than both CS-DS and CS-NT atlases compared to IM, suggesting there are more structural differences between the DSG atlas and the individual Freesurfer segmentations than there was in either CS-DS or CS-NT atlases. Upon further interrogation, we find the structural differences are predominantly on the region borders (supplemental material S1) and that differences in the DEM group (Figure 3, panel C) are likely due to brain atrophy resulting in the DSG atlas being less consistent with each individual IM segmentation.

### 3.2 PET Region of Interest (ROI) Comparisons

#### 3.2.1 DSG vs. IM Atlases

Given the structural differences found in section 3.1, we suspect such differences could impact our PET regional averages and potentially result in false positive or negative inferences about disease progression relative to those made when using IM atlases. To evaluate differences in regional PET measurements between the DSG and IM atlases, we used fixed effects linear regression. Participants’ data within a 99.8% confidence interval (+/-3 standard deviations from the mean) were included in the analysis and reported. Data outside of this range were deemed outliers and not analyzed. Results of the overall model are reported in Table 2 for regions that were significant at the p<0.05 uncorrected threshold. Prior to *p*-value adjustments, the dorsal striatum, lateral orbitofrontal cortex, and anterior cingulate were significantly different in PET ROI mean value between atlases. After adjusting the *p*-values for multiple comparisons, the dorsal striatum remained significant.

**Table 2:** Overall model statistics showing the relationship between the difference in amyloid between the atlases and the average amyloid in each region of interest, significant at p<0.05 unadjusted. Reported in the table are the brain region (i.e. ROI), *F*-value statistics, lower limit, upper limit, Standard Error (SE), pvalue, and adjusted *p*-value respectively for significant regions surviving the unadjusted p<0.05 significance threshold.

| DSG Atlas vs. IM |  |  |  |  |  |  |
| --- | --- | --- | --- | --- | --- | --- |
|  | Overall Model |  |  |  |  |  |
| Region | *F*(5,72) | Lower | Upper | *SE* | *p* | *p*<sub>adj.</sub> |
| Dorsal Striatum | 6.965 | -0.098 | 0.101 | 0.033 | 2.348e-05 | 3.520e-04 |
| Lateral Orbitofrontal | 3.261 | -0.148 | 0.164 | 0.058 | .010 | .140 |
| Anterior Cingulate | 2.470 | -0.077 | 0.136 | 0.040 | .040 | .480 |

Results for each significant ROI were visualized using Bland Altman plots (Giavarina, 2015); Figure 4). Regression lines for each diagnosis are shown as a function of the difference (DSG-IM) between the PET ROI averages computed using each atlas (y-axis) and the average amyloid load across both atlases (x-axis). Differences that are closer to zero on the y-axis indicate better correspondence between the average regional amyloid sampled from both atlases. Regression lines with positive slope indicate an increasing difference between the two atlases as a function of higher amyloid load whereas a regression line with zero slope and zero difference (y-axis) would indicate perfect correspondence across the range of amyloid between both atlases.

**Figure 4:**
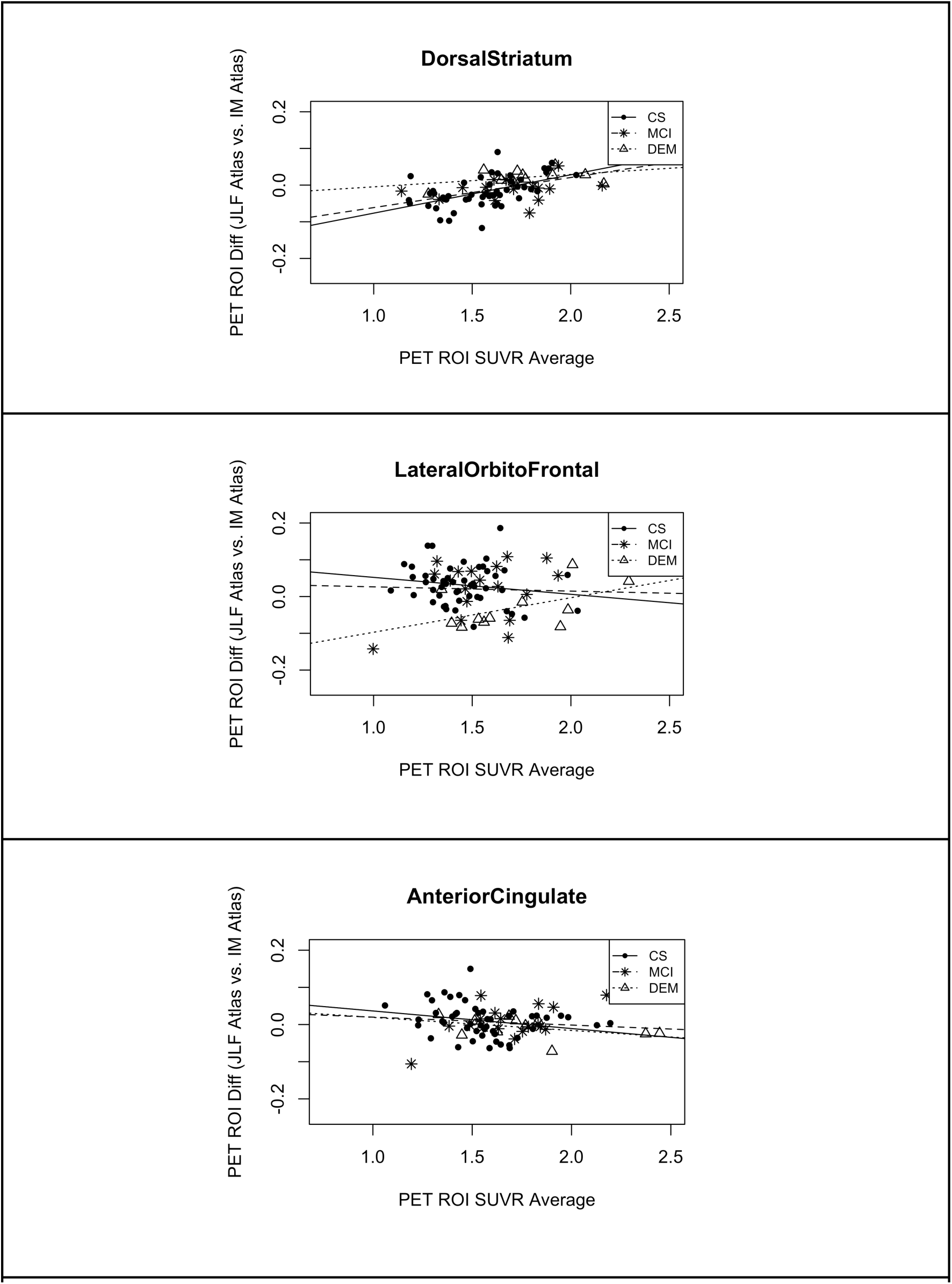
Bland Altman plots of significant regional differences in average amyloid between DSG and IM atlases. Each figure illustrates the relationship between the difference in amyloid measurements (DSG-IM) on the y-axis and the average of amyloid load on the xaxis. Regression lines with positive slope indicate an increasing difference between the two atlases as a function of higher amyloid.

#### 3.2.2 PET ROI Comparisons CS-DS vs. IM Atlases

The results from section 3.2.1 suggest that regional PET amyloid measures are reasonably consistent between the DSG and IM atlases with the exception of a few regions. Here we evaluate whether we need to account for disease status by comparing an atlas created from the cognitively-stable participants with DS (CS-DS) and sampling regional PET averages in participants with a diagnosis of MCI-DS or DEM. We expect that due to disease-related atrophy and comorbidities, using a group atlas based on those who are cognitively stable, without a more advanced form of dementia, would result in a less consistent match with the ground truth IM atlases compared to the DSG atlas. The analysis was performed using the same approach as the comparisons in 3.2.1. Results from the overall model that were significant at the p<0.05 unadjusted threshold are shown in Table 3 and the corresponding Bland Altman plots in Figure 5. Significant regions included the dorsal striatum, inferior temporal, middle temporal, and lateral orbitofrontal cortex. Similar to the DSG atlas results, dorsal striatum was the only region that remained significant after *p*-value adjustment; although, in this analysis we see temporal regions, known to be among the most affected by disease-related atrophy in Alzheimer’s disease, significant before adjustment. In contrast, the Bland Altman plots (Figure 5) show reasonably consistent regression lines between the MCI-DS and DEM groups suggesting it may not be necessary to use atlases that account for disease severity.

**Table 3:** Overall model statistics reported showing the interaction between the difference in amyloid load and the average of amyloid load in each region of interest as using IM vs. CS-DS atlas. Reported in the table are the brain region (i.e. ROI), F-value statistics, lower limit, upper limit, Standard Error (SE), *p*-value, and adjusted *p*-value respectively for significant regions surviving the unadjusted p<0.05 significance threshold..

| CS-DS Atlas vs. IM |  |  |  |  |  |  |
| --- | --- | --- | --- | --- | --- | --- |
|  | Overall Model |  |  |  |  |  |
| Region | *F*(3,25) | Lower | Upper | *SE* | *p* | *p*<sub>adj.</sub> |
| Dorsal Striatum | 8.382 | -0.035 | 0.042 | 0.017 | .001 | .015 |
| Inferior Temporal | 4.598 | -0.067 | 0.055 | 0.031 | .011 | .143 |
| Middle Temporal | 3.999 | -0.065 | 0.060 | 0.032 | .019 | .247 |
| Lateral Orbitofrontal | 3.157 | -0.113 | 0.063 | 0.045 | .042 | .504 |

**Figure 5:**
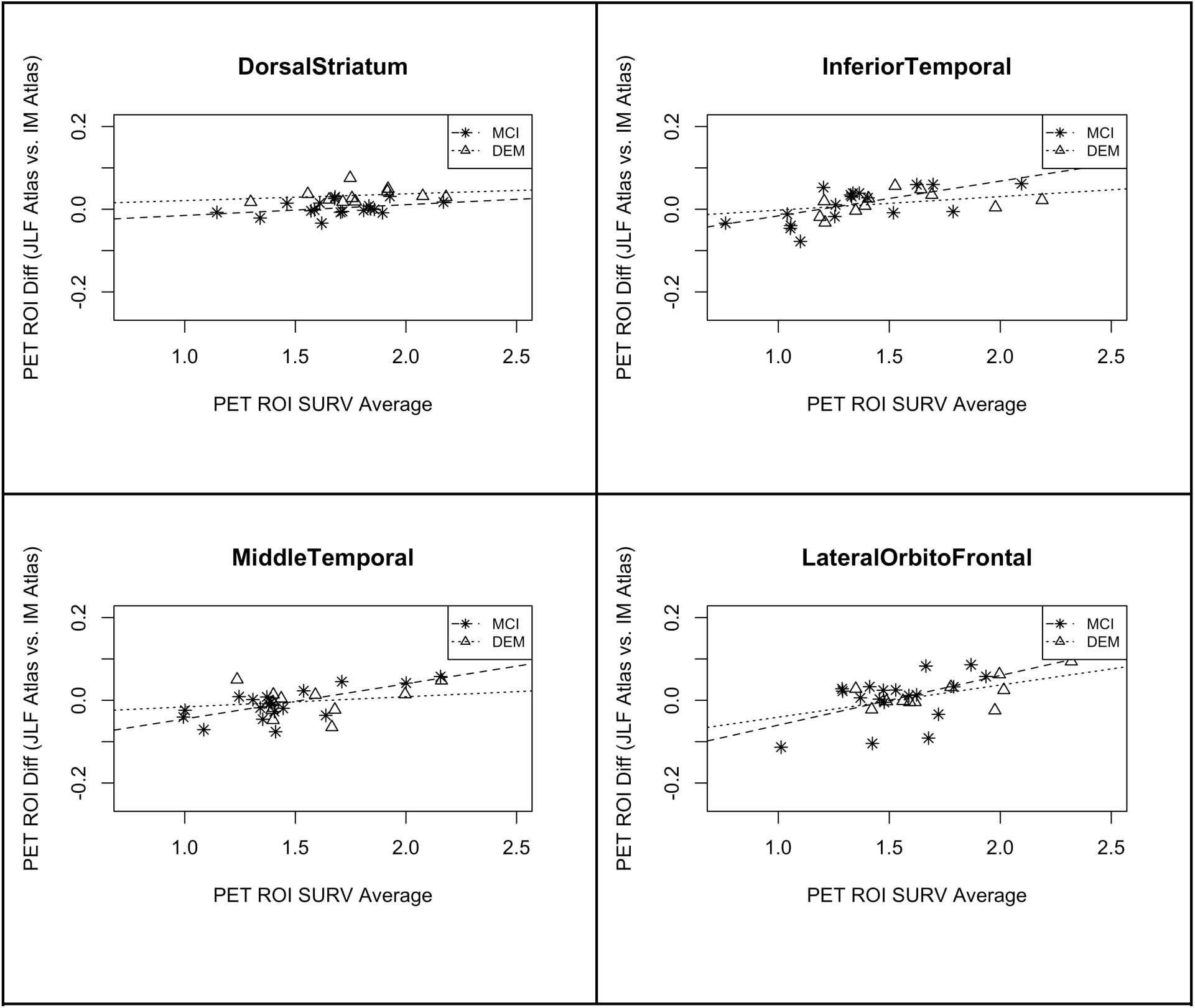
Bland Altman plots of significant regional differences in average amyloid between CS-DS and IM atlases. Each figure illustrates the relationship between the difference in amyloid measurements (CS-DS - IM) on the y-axis and the average of amyloid load on the x-axis. Regression lines with positive slope indicate an increasing difference between the two atlases as a function of higher amyloid.

#### 3.2.3 PET ROI Comparisons CS-NT vs. IM Atlases

Expanding upon the results from sections 3.2.1 and 3.2.2 which evaluated atlases made from participants with DS, with/without accounting for disease state, we now run similar experiments using an atlas made from cognitively-stable neurotypical participants of similar age. Due to brain morphological differences in participants with DS, we expect using a group atlas based on neurotypical participants will be less consistent in regional amyloid measures compared to our ground truth IM atlases. Results from the overall model that were significant at p<0.05 unadjusted are shown in Table 4 and the corresponding Bland Altman plots in Figure 6. Interestingly, similar regions to both the DSG and CS-DS atlas comparisons were identified as different from the IM atlas at the p<0.05 unadjusted threshold: dorsal striatum, middle temporal, inferior temporal along with an additional region: entorhinal cortex. Further, two additional regions approached significance: superior temporal and hippocampus. After *p*-value adjustments both the dorsal striatum and middle temporal lobe remained significant, with the entorhinal cortex approaching an adjusted significance level of p<0.05.

**Table 4:** overall model statistics reported showing the interaction between the difference in amyloid load and the average in amyloid load in each region of interest (ROI) as using IM vs. CS-NT atlas. Reported in the table are the brain region (i.e. ROI), *F*-value statistics, lower limit, upper limit, Standard Error (SE), *p*-value, and adjusted *p*-value respectively for significant regions surviving the unadjusted p<0.05 significance threshold.

| CS-NT Atlas vs. IM |  |  |  |  |  |  |
| --- | --- | --- | --- | --- | --- | --- |
|  | Overall Model |  |  |  |  |  |
| Region | *F*(5,73) | Lower | Upper | *SE* | *p* | *p*<sub>adj.</sub> |
| Dorsal Striatum | 5.611 | -0.117 | 0.089 | 0.030 | 1.979e-04 | .003 |
| Middle Temporal | 4.987 | -0.074 | 0.071 | 0.030 | .001 | .014 |
| Entorhinal Cortex | 3.863 | -0.096 | 0.175 | 0.049 | .004 | .052 |
| Inferior Temporal | 3.185 | -0.106 | 0.076 | 0.033 | .012 | .144 |

**Figure 6:**
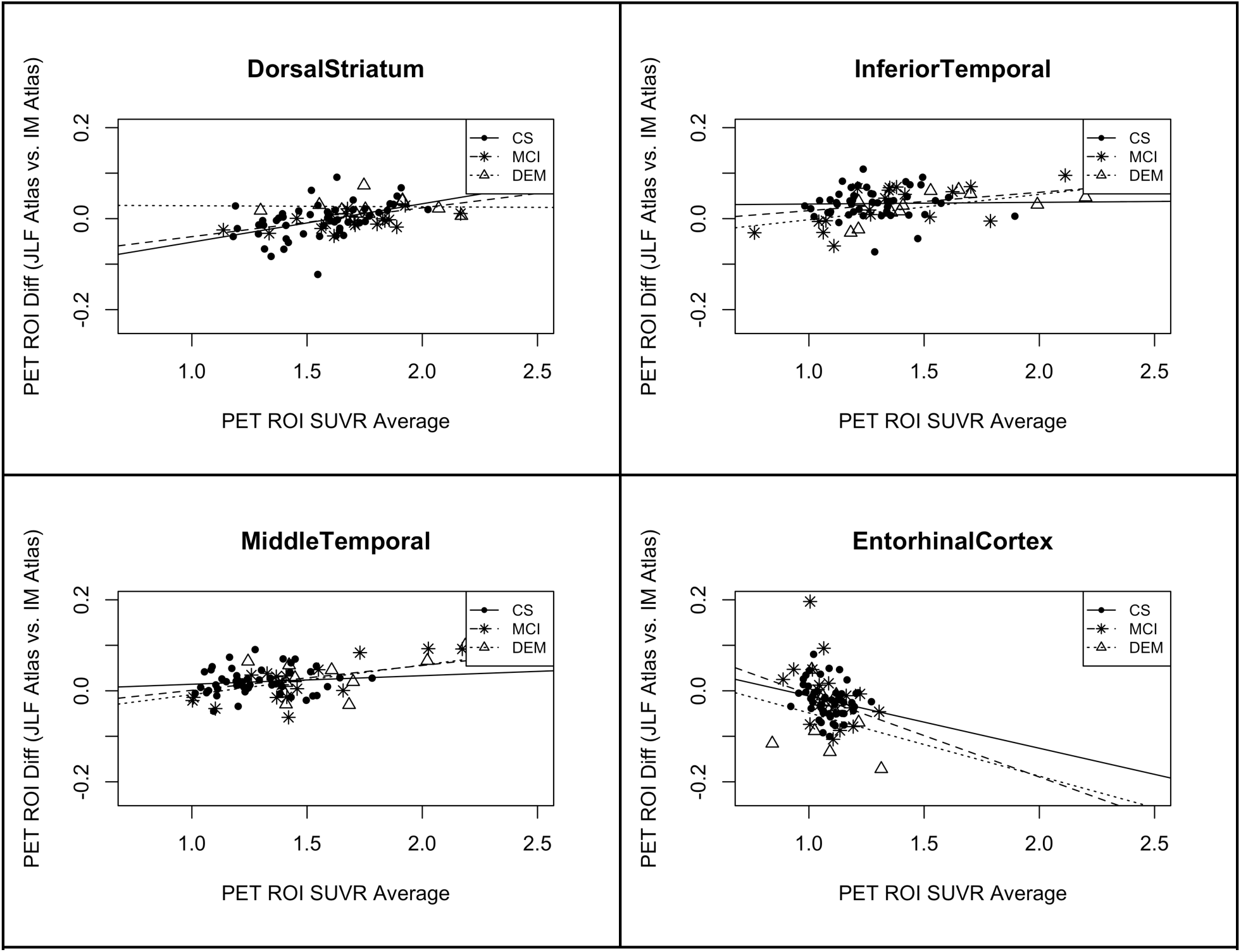
Bland Altman plots of significant regional differences in average amyloid between CS-NT and IM atlases. Each figure illustrates the relationship between the difference in amyloid measurements (CS-NT - IM) on the y-axis and the average of amyloid load on the x-axis. Regression lines with positive slope indicate an increasing difference between the two atlases as a function of higher amyloid.

### 3.3 Amyloid by Diagnostic Group Comparisons

In sections 3.1-3.2 we evaluated differences in atlas structure and regional amyloid average values, across our group atlases and the ground truth Freesurfer individual segmentions. We found some differences in structure and amyloid average values but it is unclear whether these differences would result in alternative inferences about disease progression if one were to use a group atlas instead of individual Freesurfer segmentations (IM). In this section we look at this question explicitly by focusing on typical pairwise comparisons researchers might do in the course of evaluating regional amyloid as a function of disease state in DS (e.g. DEM vs. MCI-DS, MCI-DS vs. CS, DEM vs. CS). To perform these comparisons we use fixed-effects linear regression and include age, gender, and disease status as regressors against each region’s amyloid average, for each of the atlases (e.g. DSG, CS-DS, CS-NT, and IM). The results are shown in Table 5 where we only included regions for which the inferences would have changed at the p<0.05 unadjusted significance level, either false positives or negatives, compared to our ground truth IM atlases. In comparing DEM with the CS group, we find that no inferences would have changed using any of the group atlases compared to the IM atlases. In contrast, when comparing the MCI-DS and DEM groups, we see that our inference about lateral and medial orbitofrontal would change. For lateral orbitofrontal cortex, if we used a group atlas made from cognitively stable participants with DS (CS-DS) or neurotypicals (i.e. without considering disease status; CS-NT), we would infer amyloid load is significantly different between those with a diagnosis of MCI-DS vs. DEM; whereas, if we used the DSG atlas our inference would be the same as with IM. For medial orbitofrontal cortex, we see a change when using any of the alternative group atlases, going from non-significant with IM to significant with the other atlases. In comparing the MCI-DS and CS groups we see many inferences that would have changed. Specifically, amyloid load in lateral occipital, rostral middle frontal, prefrontal cortex, medial orbitofrontal cortex, dorsal striatum, and anterior cingulate were all significantly different between the two groups using the IM atlases yet mostly (exception: rostral middle frontal using CS-DS atlas) became non-significant when using any of the group atlases. Further, the hippocampus was non-significant with the IM atlas but became significant using the group atlases.

**Table 5:**
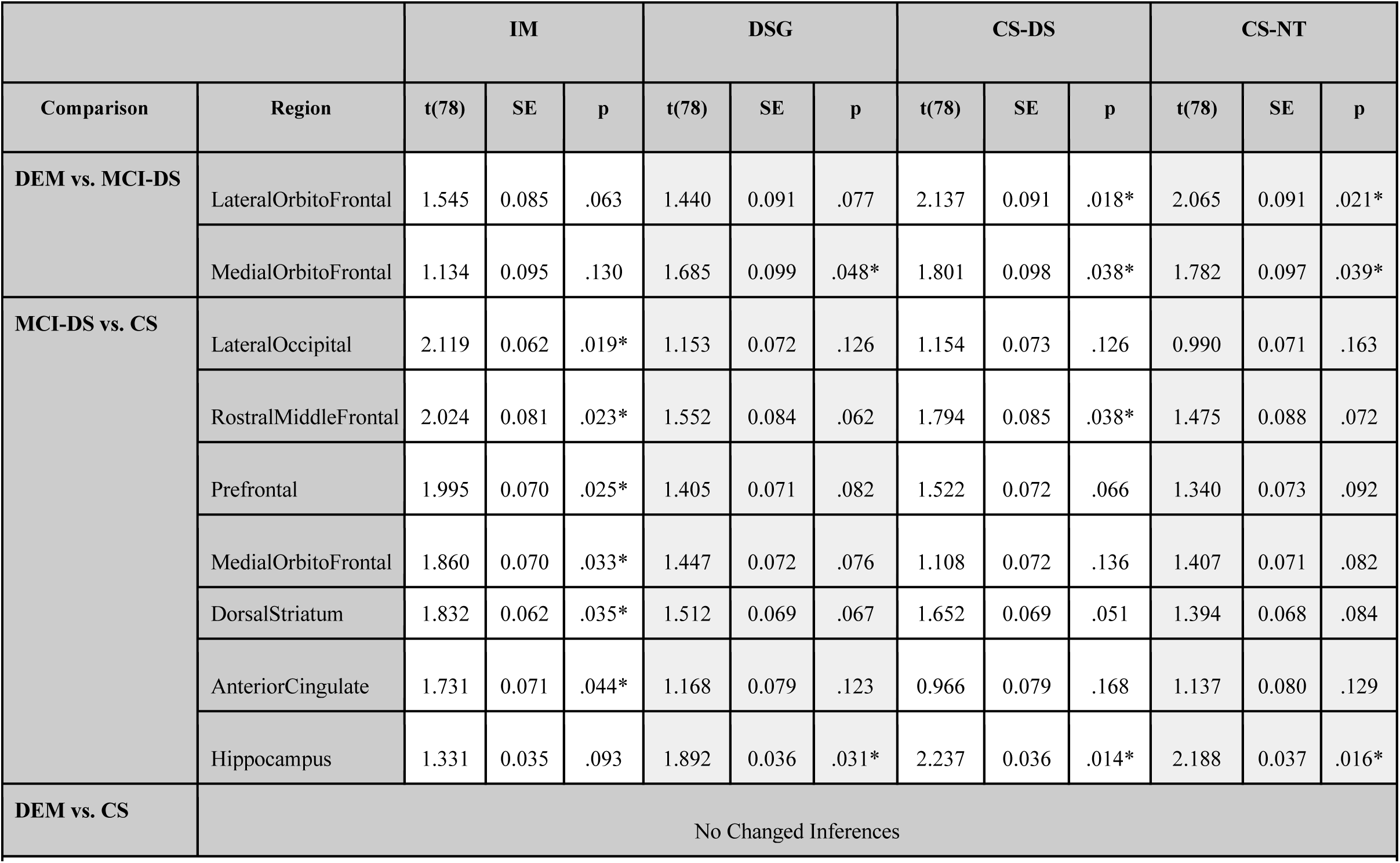
Group amyloid comparisons by region and atlas. Regions are included if inferences changed from IM when using a group atlas. Regions with significant differences surviving the p<0.05 unadjusted threshold are indicated by ‘*’. Reported in the table is the group contrast, brain region (i.e. ROI), t-statistic (t), standard error (SE), p-value (p), and adjusted p-value (p_adj_.) respectively.

## 4. DISCUSSION

In this study we created a series of probabilistic group atlases (i.e. disease-state-specific - Down Syndrome (DSG), cognitively stable - Down Syndrome (CS-DS), and cognitively stable - neurotypical (CS-NT)) and evaluated their accuracy compared to Freesurfer individual MRI segmentations (IM) when measuring regional amyloid. We hypothesized that using disease state and diagnosis-specific group (DSG) atlases would best match the ground truth IM atlas measurements, followed by CS-DS, which accounts for disorder-specific differences in brain morphology. In evaluating the structural differences in the group atlases compared to the IM atlases, we found no differences across the MCI-DS and CS groups; whereas, the DEM group had higher MSE values using the DSG atlas driven primarily by differences in each anatomical region’s boundaries.

With respect to regional amyloid measurements, we found, generally, that both the DSG and CS-DS atlases did quite well in matching IM atlas measurements with exception of the few regions identified (section 3.2). As expected, we found increasingly more differences from the IM atlases as we moved from using diagnosis and disease-state specific atlases (DSG) to disorder-specific (CS-DS) and finally the neurotypical atlas (CS-NT). For practical use, differences in regional amyloid magnitude is not necessarily significant with respect to understanding disease progression if one’s inference about differences in amyloid between cohorts doesn’t change. We therefore evaluated comparisons across the three diagnostic groups when using each of the atlases, focusing on instances when our interpretation of the differences would have changed if we were using the group atlases (section 3.3). In these comparisons we found quite a few instances where one might implicate different regions in AD disease progression if using group atlases. These effects were confined to diagnoses that were adjacent to one another in terms of disease progression (e.g. MCI-DS vs. CS, DEM vs. MCI-DS) and thus are likely to have more overlap in their amyloid distributions and smaller differences across groups. Since the two diagnostic groups (i.e. MCI-DS and CS) are adjacent, we suspect the CS sample may have included individuals with regional amyloid distributions that are similar to those in the cohort with an MCI-DS diagnosis but had not yet experienced sufficient symptoms to satisfy the criteria for MCI-DS. One limitation to note is that it’s unclear whether Freesurfer IM segmentations are capturing regional boundaries more accurately than the joint-label-fusion atlases. We suspect Freesurfer is better able to capture individual variability in anatomical region boundaries because they are based on segmenting an individual’s anatomical MRI but there may be additional variability introduced through the complex optimizations performed by Freesurfer in detecting these boundaries. Generally, the use of group and/or standardized atlases are thought to reduce subject-to-subject variability compared to individual segmentations (Thompson et al., 2000). Unlike typical Freesurfer IM segmentations that rely on prior information obtained through regression coefficient predictions and high-dimensional Bayesian optimizations (Chen et al., 2019), our group atlases use identical atlas regions (e.g. hippocampus) warped to reflect the anatomy of the participant.

In looking more closely at the regions that resulted in alternate inference in the transition from CS to MCI-DS, we observed that the dorsal striatum changed in significance when using the group atlases in addition to being the only region to survive adjusted p-value thresholds in the regional PET amyloid results (section 3.2). Autopsy studies suggest early amyloid binding in the dorsal striatum, in some cases earlier than binding in the temporal cortex, foreshadows the onset of dementia in the CS and MCI-DS diagnostic groups (Abrahamson et al., 2019; Cardemil, 2017; Keator et al., 2020b; Klunk et al., 2007). Further, there is evidence suggesting brain atrophy in striatal regions in participants with DS are consistent with findings from AD studies in neurotypical populations (Annus et al., 2017). In addition to autopsy studies, in-vivo human neuroimaging studies in DS have also shown amyloid in the striatum around 40 years of age (Handen et al., 2012; Hartley et al., 2014). Since regional amyloid retention positively correlates with regional cell loss and brain atrophy (Teipel and Hampel, 2006), we expect the early amyloid load and structural atrophy in the dorsal striatum are associated with increased variability across our participants in this region relative to other brain structures. Additional studies are needed to investigate the association between early amyloid binding and atrophy in the dorsal striatum and the impact of this association on trajectories of AD.

In summary, both the disease specific and diagnosis specific group atlases (i.e. DSG and CS-DS) performed better than the neurotypical group atlas (CS-NT) in generally replicating the regional amyloid measurements obtained from the Freesurfer individual segmentations. Overall we suggest using the CS-DS atlas as it appears to be as accurate as the DSG atlas for use in this population and better captures the overall variability in atrophy across the diagnostic groups. Each of the atlases (DSG, CS-DS, and CS-NT) along with the posterior probability maps for each region of interest (ROI) from the joint-label-fusion algorithm are made publicly available at **XXXX**. The Down Syndrome dataset used for this study is made available by the ABC-DS consortium (Crawford, n.d.). All source code for this project has been made available in a GitHub repository **(XXXX)**. Future work will focus on longitudinal group atlases to better understand the effects of disease-related brain changes on the accuracy of group atlas-based measurements and on building group atlases of younger cohorts with Down Syndrome.

## Data Availability

The down Syndrome datasets used for this study is made available by the ABC-DS consortium (Crawford, n.d.)

## Acknowledgements

The preparation of this manuscript was made possible from data obtained by the Alzheimer’s Disease in Down Syndrome (ADDS) component of the Alzheimer’s Biomarkers Consortium –Down Syndrome (ABC-DS), a longitudinal study of Alzheimer Disease biomarkers in adults with Down syndrome is supported by grants from the National Institute on Aging (NIA) (U01AG051412-01 Schupf, Lott, Silverman) and Eunice Kennedy Shriver National Institute of Child Health and Human Development (NICHD). The Co-Principal Investigators of the ADDS component of the ABC-DS program are Nicole Schupf, PhD, Dr PH. (Columbia University), Ira Lott, MD and Wayne Silverman, Ph.D. (UC Irvine). Co-Principal Investigators of the NiAD ABC-DS component are Benjamin Handen, PhD and William Klunk, MD, PhD, (University of Pittsburgh), and Bradley Christian, PhD (University of Wisconsin-Madison) Neurotypical data was collected under the Neuroimaging biomarkers for cognitive decline in elderly with amyloid pathology (NIA) grant from the National Institute of Aging (R01AG053555 Yassa, Gillen).

## References

Abrahamson, E.E., Head, E., Lott, I.T., Handen, B.L., Mufson, E.J., Christian, B.T., Klunk, W.E., Ikonomovic, M.D., 2019. Neuropathological correlates of amyloid PET imaging in Down syndrome. Dev. Neurobiol. 79, 750–766.

ADNI Home [WWW Document], n.d. URL http://www.adni-info.org/ (accessed 12.8.16).

Ali, A., Scior, K., Ratti, V., Strydom, A., King, M., Hassiotis, A., 2013. Discrimination and other barriers to accessing health care: perspectives of patients with mild and moderate intellectual disability and their carers. PLoS One 8, e70855.

Annus, T., Wilson, L.R., Acosta-Cabronero, J., Cardenas-Blanco, A., Hong, Y.T., Fryer, T.D., Coles, J.P., Menon, D.K., Zaman, S.H., Holland, A.J., Nestor, P.J., 2017. The Down syndrome brain in the presence and absence of fibrillar β-amyloidosis. Neurobiol. Aging 53, 11–19.

Annus, T., Wilson, L.R., Hong, Y.T., Acosta-Cabronero, J., Fryer, T.D., Cardenas-Blanco, A., Smith, R., Boros, I., Coles, J.P., Aigbirhio, F.I., Menon, D.K., Zaman, S.H., Nestor, P.J., Holland, A.J., 2016. The pattern of amyloid accumulation in the brains of adults with Down syndrome. Alzheimers. Dement. 12, 538–545.

Avants, B.B., Tustison, N.J., Song, G., Cook, P.A., Klein, A., Gee, J.C., 2011. A reproducible evaluation of ANTs similarity metric performance in brain image registration. Neuroimage 54, 2033–2044.

Ballard, C., Mobley, W., Hardy, J., Williams, G., Corbett, A., 2016. Dementia in Down’s syndrome. Lancet Neurol. 15, 622–636.

Bland, J.M., Martin Bland, J., Altman, D., 1986. STATISTICAL METHODS FOR ASSESSING AGREEMENT BETWEEN TWO METHODS OF CLINICAL MEASUREMENT. The Lancet. https://doi.org/10.1016/s0140-6736(86)90837-8

Brockmeyer, D., 1999. Down Syndrome and Craniovertebral Instability. Pediatric Neurosurgery. https://doi.org/10.1159/000028837

Cardemil, F., 2017. Comparison analysis and applications of the Bland-Altman method: correlation or agreement? Medwave. https://doi.org/10.5867/medwave.2016.01.6852

Chen, G., Xiao, Y., Taylor, P.A., Rajendra, J.K., Riggins, T., Geng, F., Redcay, E., Cox, R.W., 2019. Handling Multiplicity in Neuroimaging Through Bayesian Lenses with Multilevel Modeling. Neuroinformatics 17, 515–545.

Cole, J.H., Annus, T., Wilson, L.R., Remtulla, R., Hong, Y.T., Fryer, T.D., Acosta-Cabronero, J., Cardenas-Blanco, A., Smith, R., Menon, D.K., Zaman, S.H., Nestor, P.J., Holland, A.J., 2017. Brain-predicted age in Down syndrome is associated with beta amyloid deposition and cognitive decline. Neurobiol. Aging 56, 41–49.

Crawford, n.d. LONI Image Data Archive (IDA) [WWW Document]. URL https://ida.loni.usc.edu/login.jsp?project=abcds x(accessed 11.2.20).

Desikan, R.S., Ségonne, F., Fischl, B., Quinn, B.T., Dickerson, B.C., Blacker, D., Buckner, R.L., Dale, A.M., Maguire, R.P., Hyman, B.T., Albert, M.S., Killiany, R.J., 2006. An automated labeling system for subdividing the human cerebral cortex on MRI scans into gyral based regions of interest. Neuroimage 31, 968–980.

Devinsky, O., Sato, S., Conwit, R.A., Schapiro, M.B., 1990. Relation of EEG alpha background to cognitive function, brain atrophy, and cerebral metabolism in Down’s syndrome. Age-specific changes. Arch. Neurol. 47, 58–62.

Fischl, B., Salat, D.H., Busa, E., Albert, M., Dieterich, M., Haselgrove, C., van der Kouwe, A., Killiany, R., Kennedy, D., Klaveness, S., Montillo, A., Makris, N., Rosen, B., Dale, A.M., 2002. Whole brain segmentation: automated labeling of neuroanatomical structures in the human brain. Neuron 33, 341–355.

Fischl, B., van der Kouwe, A., Destrieux, C., Halgren, E., Ségonne, F., Salat, D.H., Busa, E., Seidman, L.J., Goldstein, J., Kennedy, D., Caviness, V., Makris, N., Rosen, B., Dale, A.M., 2004. Automatically parcellating the human cerebral cortex. Cereb. Cortex 14, 11–22.

FreeSurfer [WWW Document], n.d. URL https://surfer.nmr.mgh.harvard.edu/ (accessed 12.8.16).

Gardiner, K., Herault, Y., Lott, I.T., Antonarakis, S.E., Reeves, R.H., Dierssen, M., 2010. Down syndrome: from understanding the neurobiology to therapy. J. Neurosci. 30, 14943–14945.

Giavarina, D., 2015. Understanding Bland Altman analysis. Biochemia Medica. https://doi.org/10.11613/bm.2015.015

Handen, B.L., Cohen, A.D., Channamalappa, U., Bulova, P., Cannon, S.A., Cohen, W.I., Mathis, C.A., Price, J.C., Klunk, W.E., 2012. Imaging brain amyloid in nondemented young adults with Down syndrome using Pittsburgh compound B. Alzheimers. Dement. 8, 496–501.

Hartley, S.L., Handen, B.L., Devenny, D.A., Hardison, R., Mihaila, I., Price, J.C., Cohen, A.D., Klunk, W.E., Mailick, M.R., Johnson, S.C., Christian, B.T., 2014. Cognitive functioning in relation to brain amyloid-β in healthy adults with Down syndrome. Brain 137, 2556–2563.

Hommel, G., 1988. A stagewise rejective multiple test procedure based on a modified Bonferroni test. Biometrika. https://doi.org/10.1093/biomet/75.2.383

Keator, D.B., Doran, E., Taylor, L.A., Phelan, M.J., Hom, C., Tseung, K., van Erp, T.G.M., Potkin, S.G., Brickman, A.M., Rosas, D.H., Yassa, M.A., Silverman, W., Lott, I.T., 2020a. Brain Amyloid and the Transition to Dementia in Down Syndrome. Alzheimer’s & Dementia: Diagnosis, Assessment & Disease Monitoring. https://doi.org/10.1002/dad2.12126

Keator, D.B., Phelan, M.J., Taylor, L., Doran, E., Krinsky-McHale, S., Price, J., Ballard, E.E., Kreisl, W.C., Hom, C., Nguyen, D., Pulsifer, M., Lai, F., Rosas, D.H., Brickman, A.M., Schupf, N., Yassa, M.A., Silverman, W., Lott, I.T., 2020b. Down syndrome: Distribution of brain amyloid in mild cognitive impairment. Alzheimers. Dement. 12, e12013.

Klunk, W.E., Price, J.C., Mathis, C.A., Tsopelas, N.D., Lopresti, B.J., Ziolko, S.K., Bi, W., Hoge, J.A., Cohen, A.D., Ikonomovic, M.D., Saxton, J.A., Snitz, B.E., Pollen, D.A., Moonis, M., Lippa, C.F., Swearer, J.M., Johnson, K.A., Rentz, D.M., Fischman, A.J., Aizenstein, H.J., DeKosky, S.T., 2007. Amyloid deposition begins in the striatum of presenilin-1 mutation carriers from two unrelated pedigrees. J. Neurosci. 27, 6174–6184.

Lai, F., Williams, R.S., 1989. A prospective study of Alzheimer disease in Down syndrome. Arch. Neurol. 46, 849–853.

Landau, S.M., Breault, C., Joshi, A.D., Pontecorvo, M., Mathis, C.A., Jagust, W.J., Mintun, M.A., 2012. Amyloid-Imaging with Pittsburgh Compound B and Florbetapir: Comparing Radiotracers and Quantification Methods. J. Nucl. Med. 54, 70–77.

Lee, N.R., Adeyemi, E.I., Lin, A., Clasen, L.S., Lalonde, F.M., Condon, E., Driver, D.I., Shaw, P., Gogtay, N., Raznahan, A., Giedd, J.N., 2016. Dissociations in Cortical Morphometry in Youth with Down Syndrome: Evidence for Reduced Surface Area but Increased Thickness. Cereb. Cortex 26, 2982–2990.

LeMay, M., Alvarez, N., 1990. The relationship between enlargement of the temporal horns of the lateral ventricles and dementia in aging patients with Down syndrome. Neuroradiology 32, 104–107.

Lott, I.T., Lai, F., 1982. Dementia in Down’s syndrome: Observations from a neurology clinic. Applied Research in Mental Retardation. https://doi.org/10.1016/0270-3092(82)90017-0

Mann, D.M., Jones, D., Prinja, D., Purkiss, M.S., 1990. The prevalence of amyloid (A4) protein deposits within the cerebral and cerebellar cortex in Down’s syndrome and Alzheimer’s disease. Acta Neuropathol. 80, 318–327.

Parker, S.E., Mai, C.T., Canfield, M.A., Rickard, R., Wang, Y., Meyer, R.E., Anderson, P., Mason, C.A., Collins, J.S., Kirby, R.S., Correa, A., National Birth Defects Prevention Network, 2010. Updated National Birth Prevalence estimates for selected birth defects in the United States, 2004-2006. Birth Defects Res. A Clin. Mol. Teratol. 88, 1008–1016.

Pearlson, G.D., Breiter, S.N., Aylward, E.H., Warren, A.C., Grygorcewicz, M., Frangou, S., Barta, P.E., Pulsifer, M.B., 1998. MRI brain changes in subjects with Down syndrome with and without dementia. Dev. Med. Child Neurol. 40, 326–334.

Prasher, V., Cumella, S., Natarajan, K., Rolfe, E., Shah, S., Haque, M.S., 2003. Magnetic resonance imaging, Down’s syndrome and Alzheimer’s disease: research and clinical implications. J. Intellect. Disabil. Res. 47, 90–100.

Quigley, H., Colloby, S.J., O’Brien, J.T., 2011. PET imaging of brain amyloid in dementia: a review. Int. J. Geriatr. Psychiatry 26, 991–999.

Rabinovici, G.D., Jagust, W.J., 2009. Amyloid imaging in aging and dementia: testing the amyloid hypothesis in vivo. Behav. Neurol. 21, 117–128.

Radhakrishnan, R., Towbin, A.J., 2014. Imaging findings in Down syndrome. Pediatr. Radiol. 44, 506– 521.

Raz, N., Torres, I.J., Briggs, S.D., Spencer, W.D., Thornton, A.E., Loken, W.J., Gunning, F.M., McQuain, J.D., Driesen, N.R., Acker, J.D., 1995. Selective neuroanatomic abnormalities in Down’s syndrome and their cognitive correlates: evidence from MRI morphometry. Neurology 45, 356–366.

Rodrigues, M., Nunes, J., Figueiredo, S., Martins de Campos, A., Geraldo, A.F., 2019. Neuroimaging assessment in Down syndrome: a pictorial review. Insights Imaging 10, 52.

Silverman, W., 2007. Down syndrome: cognitive phenotype. Ment. Retard. Dev. Disabil. Res. Rev. 13, 228–236.

Silverman, W., Schupf, N., Zigman, W., Devenny, D., Miezejeski, C., Schubert, R., Ryan, R., 2004. Dementia in Adults With Mental Retardation: Assessment at a Single Point in Time. American Journal on Mental Retardation. https://doi.org/2.0.co;2”>10.1352/0895-8017(2004)109<111:diawmr>2.0.co;2

Teipel, S.J., Hampel, H., 2006. Neuroanatomy of Down Syndrome in vivo: A Model of Preclinical Alzheimer’s Disease. Behavior Genetics. https://doi.org/10.1007/s10519-006-9047-x

Thompson, P., Mega, M.S., Toga, A.W., 2000. Disease-Specific Probabilistic Brain Atlases. Proc. IEEE Comput. Soc. Conf. Comput. Vis. Pattern Recognit. 2000, 227–234.

Toga, A.W., Thompson, P.M., 2002. New Approaches in Brain Morphometry. The American Journal of Geriatric Psychiatry. https://doi.org/10.1097/00019442-200201000-00003

Weis, S., Weber, G., Neuhold, A., Rett, A., 1991. Down syndrome: MR quantification of brain structures and comparison with normal control subjects. AJNR Am. J. Neuroradiol. 12, 1207–1211.

Wullink, M., Veldhuijzen, W., Lantman-de Valk, H.M. van S., Metsemakers, J.F.M., Dinant, G.-J., 2009. Doctor-patient communication with people with intellectual disability--a qualitative study. BMC Fam. Pract. 10, 82.

