## Supplementary material for "Joint-Label Fusion Brain Atlases for Dementia Research in Down Syndrome": S1, S2 (S2.1, S2.2, S2.3)

Corresponding Author:

Nazek Queder

### Abstract

Research suggests a link between Alzheimer's Disease in Down Syndrome (DS) and the overexpression of amyloid plaques. Using Positron Emission Tomography (PET) we can assess the in-vivo regional amyloid load using several available ligands. To measure amyloid distributions in specific brain regions, a brain atlas is used. A popular method of creating a brain atlas is to segment a participant's structural Magnetic Resonance Imaging (MRI) scan. Acquiring an MRI is often challenging in intellectually-imparied populations because of contraindications or data exclusion due to significant motion artifacts or incomplete sequences related to general discomfort. When an MRI cannot be acquired, it is typically replaced with a standardized brain atlas derived from neurotypical populations which may be inappropriate for use in DS. In this project, we create a series of disease and diagnosis-specific (cognitively stable, mild cognitive impairment (MCI-DS), and dementia) probabilistic group atlases of participants with DS and evaluate their accuracy of quantifying regional amyloid load compared to our ground truth individual MRI-based segmentations. Further, we compare the diagnostic-specific atlases with a probabilistic atlas constructed from similar-aged cognitively-stable neurotypical participants. We hypothesized that regional PET signals will best match the ground truth by using DS group atlases that aligns with a participant's disorder and disease status (e.g. DS and MCI-DS). Our results vary by brain region but generally show that using a disorder-specific atlas in DS better matches the ground truth than using an atlas constructed from cognitively-stable neurotypical participants. We found no additional benefit of using a disease-state specific atlas. All atlases are made publicly available for the research community.

**Keywords:** Down Syndrome, dementia, Joint-label-fusion atlas, MRI, Amyloid PET, Region of Interest.

**Abbreviations:** AD, DS, Aβ, DSG, CS-DS, CS-NT, LOOCV, ROI, MSE, MRI, PET, JLF, CS, MCI-DS, DEM.

SUPPLEMENTAL MATERIAL:

**S1. Differences in Segmentations:**

Our study mainly focused on quantifying the differences between our atlases (i.e. DSG, CS-DS, CS-NT) and the Individual MRI (IM) Freesurfer segmentations. To understand the structural differences in segmentation labels in each region of interest (ROI), each participant's IM was contrasted with our atlases to calculate the mean square error (MSE) and to produce a Freesurfer segmented image of the difference. Shown in Figure S1, the first two columns illustrate the quantification between the IM segmentations atlas subtracted from each of our atlases (i.e. DSG, CS-DS, CS-NT) with the last column representing the difference in segments between the two atlases. Only Freesurfer segmentation images of participants who's MSE scores were identified as outliers were included in the illustration in Figure S. It's important to note that the differences are often located along the region boundaries with a few exceptions.

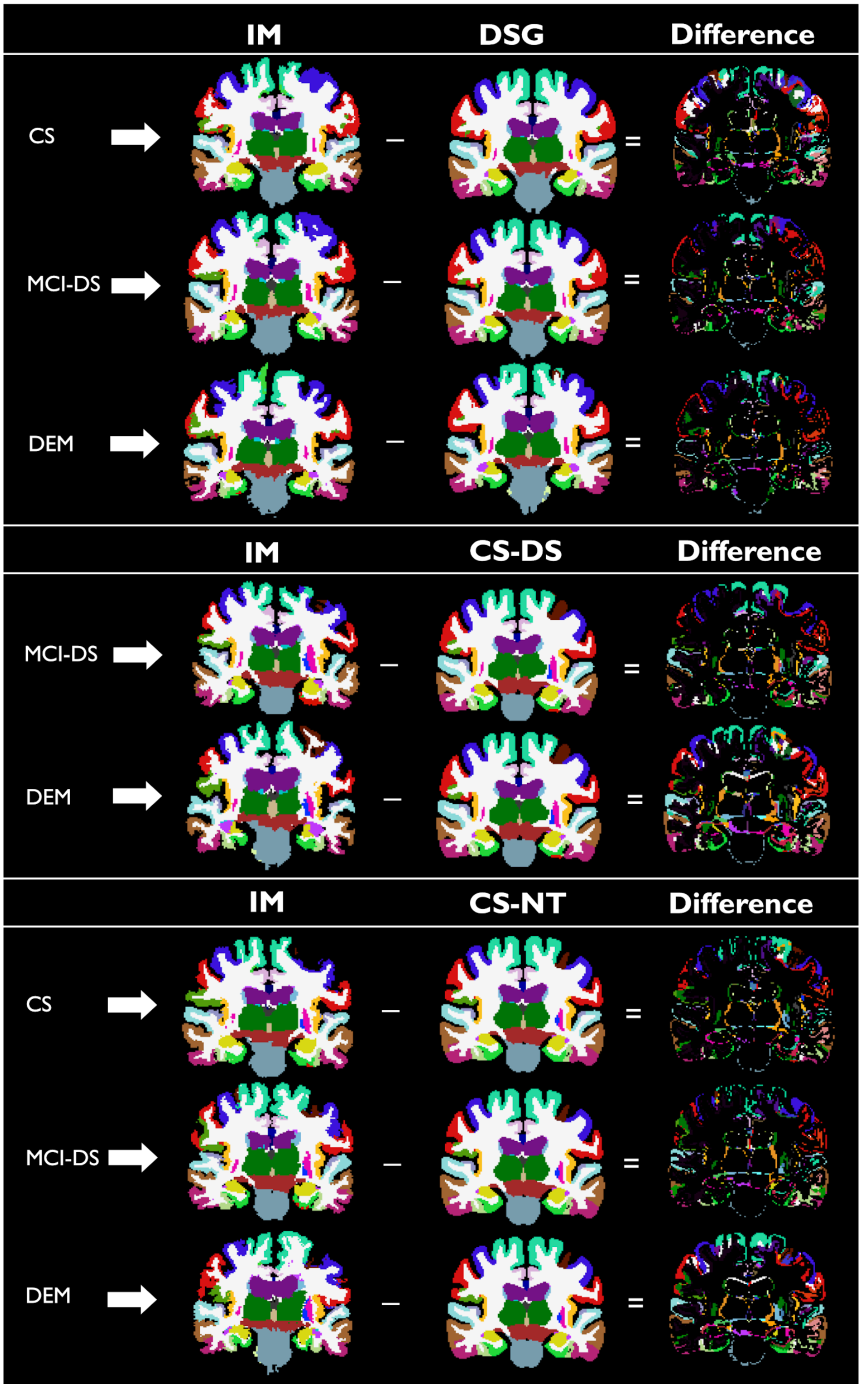


Figure S1: The top section includes examples showing three DSG atlases constructed using participants with DS and within each disease state (e.g. CS, MCI-DS, or DEM) compared to selected individual (IM) atlases of the left out participant. The middle and the last section include examples showing CS-DS and CS-NT atlases constructed from cognitively stable participants with DS and neurotypical participants respectively compared to IM atlases within disease status. The last column shows the difference between the atlases.

**S2. ROI PET Amyloid Linear Regression Analysis**

Fixed linear regression analysis was performed to evaluate the impact of regional amyloid load on the differences in PET amyloid average values between our atlases (i.e. DSG, CS-DS, CS-NT) and individual MRI (IM) segmentations, shown in figures S1.1, S1.2, and S1.3. The model evaluates the differences between our atlases and IM respectively as a function of the mean amyloid load of the two atlases in a region of interest (ROI) at $p<0.05$ significance level. Regression lines with positive estimates indicate higher PET amyloid average values given by our atlases compared to IM. Results indicate that the significance of the results varies across our atlases and the three diagnostic groups (cognitivley stabled (CS), Mild cognitive impairment - Down Syndrome (MCI-DS), and Demented (DEM)). Surviving the p-value adjustments: as the regional mean amyloid load increases, both the DSG and the CS-NT atlases resulted in higher estimation of PET amyloid average values in the Dorsal Striatum for the CS group and in the Anterior Cingulate for the MCI-DS group, whereas the CS-DS atlas gave higher estimate in the Inferior Temporal and the Middle Temporal for the MCI-DS groups. No ROI's survived the p-value adjustments for the DEM group across our atlases. The increase in the mean amyloid load between DSG and CS-NT atlases and IM impacts the differences in same regions (i.e. Dorsal Striatum and the Anterior Cingulate) across the diagnostic groups. In contrast, the DS-CS results have shown that only MCI-DS is impacted by the increase of amyloid load in both the inferior and middle temporals.

S2.1 ROI Linear Regression Model Analysis Between DSG Atlases and IM

| | CS | | | | MCI-DS | | | | DEM | | | |
|---|---|---|---|---|---|---|---|---|---|---|---|---|
| **Region** | ***Estimate*** | ***t(48)*** | ***p*** | ***Adj p*** | ***Estimate*** | ***t(17)*** | ***p*** | ***Adj p*** | ***Estimate*** | ***t(10)*** | ***p*** | ***Adj p*** |
| Anterior Cingulate | -0.047 | -1.861 | .067 | .670 | 0.155 | 3.058 | .003* | .045* | 0.012 | 0.276 | .783 | .968 |
| Dorsal Striatum | 0.105 | 4.556 | .000* | .000* | -0.080 | -1.924 | .058 | .638 | -0.071 | -1.469 | .146 | .968 |
| Entorhinal Cortex | 0.077 | 0.812 | .419 | .956 | -0.085 | -0.578 | .565 | .984 | -0.101 | -0.597 | .553 | .968 |
| Hippocampus | 0.024 | 0.479 | .634 | .956 | 0.036 | 0.505 | .615 | .984 | -0.073 | -0.887 | .378 | .968 |
| Inferior Parietal | -0.073 | -2.069 | .042* | .504 | 0.109 | 2.276 | .026* | .312 | 0.082 | 1.546 | .127 | .968 |
| Inferior Temporal | 0.018 | 0.443 | .659 | .956 | 0.047 | 0.826 | .411 | .984 | 0.062 | 0.976 | .333 | .968 |
| Lateral Orbitofrontal | -0.046 | -1.052 | .296 | .956 | 0.149 | 1.942 | .056 | .616 | 0.140 | 1.884 | .064 | .747 |
| Lateral Occipital | 0.058 | 1.196 | .236 | .956 | -0.023 | -0.341 | .734 | .984 | -0.010 | -0.140 | .889 | .968 |
| Medial Orbitofrontal | -0.038 | -1.254 | .214 | .956 | 0.001 | 0.020 | .984 | .984 | 0.087 | 1.675 | .098 | .945 |
| Middle Temporal | -0.002 | -0.055 | .956 | .956 | 0.051 | 1.256 | .213 | .984 | 0.060 | 1.261 | .211 | .968 |
| Orbitofrontal | -0.036 | -1.060 | .293 | .956 | 0.085 | 1.395 | .167 | .984 | 0.123 | 2.148 | .035* | .490 |
| Posterior Cingulate | -0.029 | -1.612 | .111 | .888 | 0.019 | 0.535 | .594 | .984 | 0.012 | 0.360 | .720 | .968 |
| Rostral Middle Frontal | -0.005 | -0.164 | .870 | .956 | -0.028 | -0.583 | .562 | .984 | -0.002 | -0.040 | .968 | .968 |
| Superior Frontal | -0.051 | -1.952 | .055 | .605 | 0.086 | 1.955 | .054 | .594 | 0.057 | 1.163 | .249 | .968 |
| Superior Temporal | -0.009 | -0.256 | .798 | .956 | -0.010 | -0.197 | .844 | .984 | -0.011 | -0.175 | .862 | .968 |

Figure S2.1: This table includes linear regression results comparing the average amyloid differences when using the DSG atlas compared to the IM atlas for each diagnostic group. Negative t-values indicate regions where the DSG atlas yields lower average amyloid compared to the IM atlas.

S2.2 ROI Linear Regression Model Analysis Between CS-DS Atlas and IM

| | MCI-DS | | | | DEM | | | |
|---|---|---|---|---|---|---|---|---|
| **Region** | ***Estimate*** | ***t(17)*** | ***p*** | ***Adj p*** | ***Estimate*** | ***t(10)*** | ***p*** | ***Adj p*** |
| Anterior Cingulate | 0.090 | 2.244 | .034* | .408 | -0.087 | -1.689 | .104 | .985 |
| Dorsal Striatum | 0.026 | 1.463 | .156 | .849 | -0.010 | -0.339 | .737 | .985 |
| Entorhinal Cortex | -0.017 | -0.193 | .849 | .849 | 0.075 | 0.558 | .582 | .985 |
| Hippocampus | 0.033 | 0.926 | .363 | .849 | -0.026 | -0.470 | .642 | .985 |
| Inferior Parietal | 0.029 | 0.877 | .389 | .849 | -0.075 | -1.430 | .165 | .985 |
| Inferior Temporal | 0.084 | 3.520 | .002* | .028* | -0.051 | -1.328 | .196 | .985 |
| Lateral Orbitofrontal | 0.119 | 2.492 | .020* | .240 | -0.042 | -0.622 | .539 | .985 |
| Lateral Occipital | 0.076 | 1.632 | .115 | .849 | -0.049 | -0.683 | .501 | .985 |
| Medial Orbitofrontal | -0.018 | -0.324 | .749 | .849 | 0.037 | 0.490 | .628 | .985 |
| Middle Temporal | 0.085 | 3.319 | .003* | .042* | -0.060 | -1.372 | .182 | .985 |
| Orbitofrontal | 0.064 | 1.327 | .196 | .849 | -0.001 | -0.019 | .985 | .985 |
| Posterior Cingulate | -0.011 | -0.424 | .675 | .849 | 0.032 | 0.949 | .352 | .985 |
| Rostral Middle Frontal | -0.026 | -0.607 | .549 | .849 | 0.042 | 0.672 | .508 | .985 |
| Superior Frontal | 0.027 | 0.699 | .491 | .849 | 0.011 | 0.186 | .854 | .985 |
| Superior Temporal | 0.037 | 1.091 | .286 | .849 | 0.007 | 0.131 | .897 | .985 |

Figure S2.2: This table includes linear regression results comparing the average amyloid differences when using the CS-DS atlas compared to the IM atlas for each diagnostic group. Negative t-values indicate regions where the CS-DS atlas yields lower average amyloid compared to the IM atlas.

S2.3 ROI Linear Regression Model Analysis Between CS-NT Atlas and IM

| | CS | | | | MCI-DS | | | | DEM | | | |
|---|---|---|---|---|---|---|---|---|---|---|---|---|
| **Region** | ***Estimate*** | ***t(49)*** | ***p*** | ***Adj p*** | ***Estimate*** | ***t(17)*** | ***p*** | ***Adj p*** | ***Estimate*** | ***t(9)*** | ***p*** | ***Adj p*** |
| Anterior Cingulate | -0.041 | -1.646 | 0.104 | 0.885 | 0.155 | 3.107 | .003* | .045* | 0.046 | 1.074 | 0.286 | 0.931 |
| Dorsal Striatum | 0.084 | 4.026 | 1.37E-04 | .002* | -0.046 | -1.225 | 0.224 | 0.896 | -0.087 | -1.94 | .056* | 0.672 |
| Entorhinal Cortex | -0.115 | -1.233 | 0.221 | 0.885 | -0.203 | -1.37 | 0.175 | 0.81 | -0.024 | -0.15 | 0.881 | 0.931 |
| Hippocampus | 0.028 | 0.763 | 0.448 | 0.885 | 0.013 | 0.238 | 0.812 | 0.948 | -0.018 | -0.307 | 0.76 | 0.931 |
| Inferior Parietal | -0.043 | -1.101 | 0.275 | 0.885 | 0.046 | 0.849 | 0.398 | 0.948 | -0.005 | -0.087 | 0.931 | 0.931 |
| Inferior Temporal | 0.004 | 0.145 | 0.885 | 0.885 | 0.083 | 2.298 | .024* | 0.288 | 0.052 | 1.274 | 0.207 | 0.931 |
| Lateral Occipital | -0.012 | -0.285 | 0.776 | 0.885 | 0.085 | 1.421 | 0.16 | 0.8 | 0.042 | 0.635 | 0.527 | 0.931 |
| Lateral Orbitofrontal | -0.029 | -0.814 | 0.418 | 0.885 | 0.149 | 2.371 | .020* | 0.24 | 0.098 | 1.572 | 0.12 | 0.931 |
| Medial Orbitofrontal | -0.028 | -0.786 | 0.435 | 0.885 | -0.004 | -0.066 | 0.948 | 0.948 | 0.038 | 0.594 | 0.554 | 0.931 |
| Middle Temporal | 0.019 | 0.775 | 0.441 | 0.885 | 0.085 | 2.505 | .014* | 0.168 | 0.046 | 1.13 | 0.262 | 0.931 |
| Orbitofrontal | -0.018 | -0.52 | 0.605 | 0.885 | 0.077 | 1.269 | 0.209 | 0.836 | 0.071 | 1.218 | 0.227 | 0.931 |
| Posterior Cingulate | -0.027 | -1.574 | 0.12 | 0.885 | 0.024 | 0.7 | 0.486 | 0.948 | 0.041 | 1.352 | 0.18 | 0.931 |
| Rostral Middle Frontal | 0.053 | 1.544 | 0.127 | 0.885 | -0.073 | -1.26 | 0.212 | 0.848 | -0.024 | -0.401 | 0.69 | 0.931 |
| Superior Frontal | -0.026 | -0.957 | 0.342 | 0.885 | 0.066 | 1.401 | 0.165 | 0.81 | 0.068 | 1.294 | 0.2 | 0.931 |
| Superior Temporal | 0.012 | 0.368 | 0.714 | 0.885 | 0.037 | 0.78 | 0.438 | 0.948 | 0.043 | 0.767 | 0.446 | 0.931 |

Figure S2.3: This table includes linear regression results comparing the average amyloid differences when using the CS-NT atlas compared to the IM atlas for each diagnostic group. Negative t-values indicate regions where the CS-NT atlas yields lower average amyloid compared to the IM atlas.
